# Endothelin-1 is Increased in the Plasma of Patients Hospitalized with Covid-19

**DOI:** 10.1101/2021.12.30.21268236

**Authors:** George R. Abraham, Rhoda E. Kuc, Magnus Althage, Peter J. Greasley, Philip Ambery, Janet J. Maguire, Ian B. Wilkinson, Stephen P. Hoole, Joseph Cheriyan, Anthony P. Davenport

## Abstract

**Importance:** The coronavirus disease 2019 (Covid-19) pandemic continues to place a devastating strain on healthcare services worldwide and there remains an ongoing requirement for new treatments. A key mechanism recognised in progressive severe disease is virus-induced endothelial dysregulation. Endothelin-1 (ET-1), being the most highly expressed peptide in endothelial cells and potent vasoconstrictor of human blood vessels, represents a potential therapeutic target through the use of Endothelin receptor antagonists.

**Objective:** To investigate the association of plasma ET-1 with Covid-19 disease severity

**Design:** Retrospective longitudinal cohort study of Covid-19 patients divided into Group A (asymptomatic or symptoms not requiring hospitalisation), Group B (symptoms requiring hospitalisation) and Group C (symptoms requiring supplemental oxygen therapy or assisted ventilation) recruited between March and July 2020 (the first wave of the Covid-19 pandemic in the UK). Data were compared with a contemporaneous cross-section of non-infected volunteers (Controls).

**Setting:** Single Tertiary National Health Service Hospital.

**Participants:** Tissue banked plasma samples were obtained from 194 patients.

**Exposures:** Quantitation of ET-1 in plasma by specific enzyme linked immunosorbent assay.

**Main outcome and measures:** Pairwise comparison of ET-1 levels (median [IQR]) between patient categories, and subgroups defined by clinical outcomes.

**Results:** Baseline ET-1 plasma levels (pg/ml) were elevated in patients requiring hospitalisation compared with controls and patients with asymptomatic or mild infection (Group B: 1.59 [1.13-1.98], and Group C: 1.65 [1.02-2.32] versus controls: 0.68 [0.47-0.87], p=<0.001 and Group A: 0.72 [0.57-1.10], p=<0.001). ET-1 levels were also elevated in patients that died (2.09 [1.66-3.15]), developed acute kidney (1.70 [1.07-2.36]) or myocardial injury (1.50 [0.92-2.28]) compared with patients with an uncomplicated infection (1.00 [0.61-1.57], p=<0.01). Amongst surviving hospitalised patients, ET-1 concentrations decreased when measured at 28 days (Group B: 0.86 [0.60-1.61] and Group C: 1.17 [0.66-1.62] versus baseline, p=<0.05) and 90 days (Group B: 0.69 [0.59-1.38] and Group C: 1.01 [0.64-1.21] versus baseline, p=<0.05).

**Conclusions and relevance:** Hospitalised Covid-19 patients demonstrate elevated ET-1 levels during the acute phase of infection and this is associated with increasing clinical severity of the disease. The results support the hypothesis that endothelin receptor antagonists may be beneficial for certain Covid-19 patients.

**Key Points:** *Question:* What is the association of the endothelial peptide and potent vasoconstrictor: endothelin-1 with disease severity in Covid-19 infection?

*Findings:* Hospitalised Covid-19 patients (especially those requiring supplemental oxygen and assisted ventilation, dying patients, and those who developed acute myocardial or kidney injury) have higher circulating endothelin-1 levels during the acute phase of their infection, compared with patients with asymptomatic or only mildly symptomatic Covid-19 infection.

*Meaning:* Endothelial dysregulation is a well-recognised mechanism for progressive severe Covid-19 infection and these results suggest targeting endothelin-1 activity through the use of Endothelin receptor antagonists may be of benefit.

## Introduction

The Coronavirus disease 2019 (Covid-19) pandemic continues to place a devastating strain on healthcare services worldwide and there remains an ongoing requirement for new therapies^1^.

There is emerging consensus that in progressive severe Covid-19 disease, virus-induced endothelial damage may result in a syndrome of excessive vasoconstriction, inflammation and thrombosis^2,3^. Viral inclusions and lymphocytic infiltration of apoptotic endothelial cells have been identified from histological examination of tissues obtained from Covid-19 patients with acute lung^4,5^, kidney^5^ and myocardial injury^5,6^. Additionally, multiple plasma markers for endothelial injury are elevated in hospitalised Covid-19 patients on admission^7,8^.

Endothelin-1 (ET-1), being the most highly expressed peptide in endothelial cells and potent vasoconstrictor of human blood vessels^9,10^, represents a potential therapeutic target. The benefit of endothelin receptor antagonists is already well established in pulmonary arterial hypertension^11^ hence these medications may be suitable for accelerated regulatory approval. ET-1 is released from endothelial cells via a continuous constitutive pathway and supplemented by ET-1 release from Weibel-Palade bodies (the unique storage granules of endothelial cells) in response to extra-cellular stimuli including inflammatory cytokines^10^. Serialised measurement of plasma ET-1 during Covid-19 infection has not previously been undertaken. We hypothesised that elevated levels of plasma ET-1 in the acute phase of Covid-19 infection would be associated with more severe disease.

## Methods

Plasma samples and clinical variables were obtained with ethical approval (REC:17/EE/0025) and informed consent from participants enrolled in the Cambridge NIHR Covid-19 Biobank project. Patients with positive SARS-Cov-2 PCR tests were divided into three groups based on the clinical severity of their illness: A, asymptomatic or mild symptoms not requiring hospitalisation; B, symptoms requiring hospitalisation but with no requirement for supplemental oxygen therapy at any stage; C, hospitalised with symptoms requiring one, or a combination of supplemental oxygen therapy, non-invasive or invasive ventilation. Patient demographics and clinical codes for underlying comorbidities (hypertension, ischemic heart disease, diabetes, congestive cardiac failure and chronic kidney disease) were retrospectively obtained from the electronic medical record as was the occurrence during the index admission of the following clinical endpoints: acute kidney injury (defined according to the Kidney Disease Improving Global Outcomes guidelines (2012): an acute rise in serum creatinine ≥26.5 μmol/l from baseline), acute myocardial injury (defined according to the Fourth Universal Definition of Myocardial Infarction (2018): an acute rise in cardiac troponin level with at least one value >99th percentile upper reference limit) and inpatient mortality. Venous blood samples were collected (where possible after the time of enrolment) from patients at 0, 28 and 90 days after admission to hospital (or from the time of PCR test for Group A). Samples were also collected at baseline for a control group comprising PCR negative hospital staff. ET-1 was measured in duplicate using an established sandwich ELISA (R&D Systems, U.S.A).

Statistical analysis was performed using SPSS version 27 (IBM Corp., USA) for Windows. Normality of continuous variable distributions was tested by one-sample Kolmogorov-Smirnov test. Non-normally distributed continuous variables are presented as Median (inter-quartile range [Q1 - Q3]). Pairwise comparison of ET-1 levels between patient categories A-C and controls, between different time-points for categories A-C, and between subgroups defined by clinical endpoints was undertaken using the Independent Samples Kruskal-Wallis Test. To assess for potential confounding effects when comparing ET-1 levels between patient categories A-C and controls, univariate analysis of co-variance was performed for each of: age, gender, hypertension, ischemic heart disease, diabetes, congestive cardiac failure and chronic kidney disease.

## Results

All data that support the findings of this study are available from the corresponding author upon reasonable request. Plasma samples were obtained from 194 patients. Samples from 157 patients were available at baseline, 84 patients at day 28 and 35 patients at day 90. Reasons for drop-out during follow-up included death, persisting disability and repatriation outside our locality, having been initially referred to our centre for specialist care. Plasma samples were sufficient for analysis in all cases. At baseline, ET-1 levels (pg/ml) were significantly elevated in the patients requiring hospitalisation (Group B: 1.59 [1.13-1.98], and C: 1.65 [1.02-2.32]) compared with both controls (0.68 [0.47-0.87], p=<0.001) and patients with asymptomatic or mild infection (Group A: 0.72 [0.57-1.10], p=<0.001): *Figure 1A*. Mean age±SD was 41.1±16.4 in controls, 35.8±12.6 in group A, 58.8±17.1 in group B and 62.1±14.3 in Group C. The proportion of females was 61.5% in controls, 82.4% in group A, 38.5% in Group B and 32.1% in Group C. The frequency of underlying comorbidities was higher in Groups B and C compared with Group A and controls (*Table 1*). Despite heterogeneity in clinical and demographic characteristics, differences in baseline ET-1 between all patient categories remained significant (p<0.05) in corrected models for age, gender, hypertension, ischemic heart disease, diabetes, congestive cardiac failure and chronic kidney disease using between-subjects effect analysis of co-variance (*supplemental information)*.

**Table 1:**
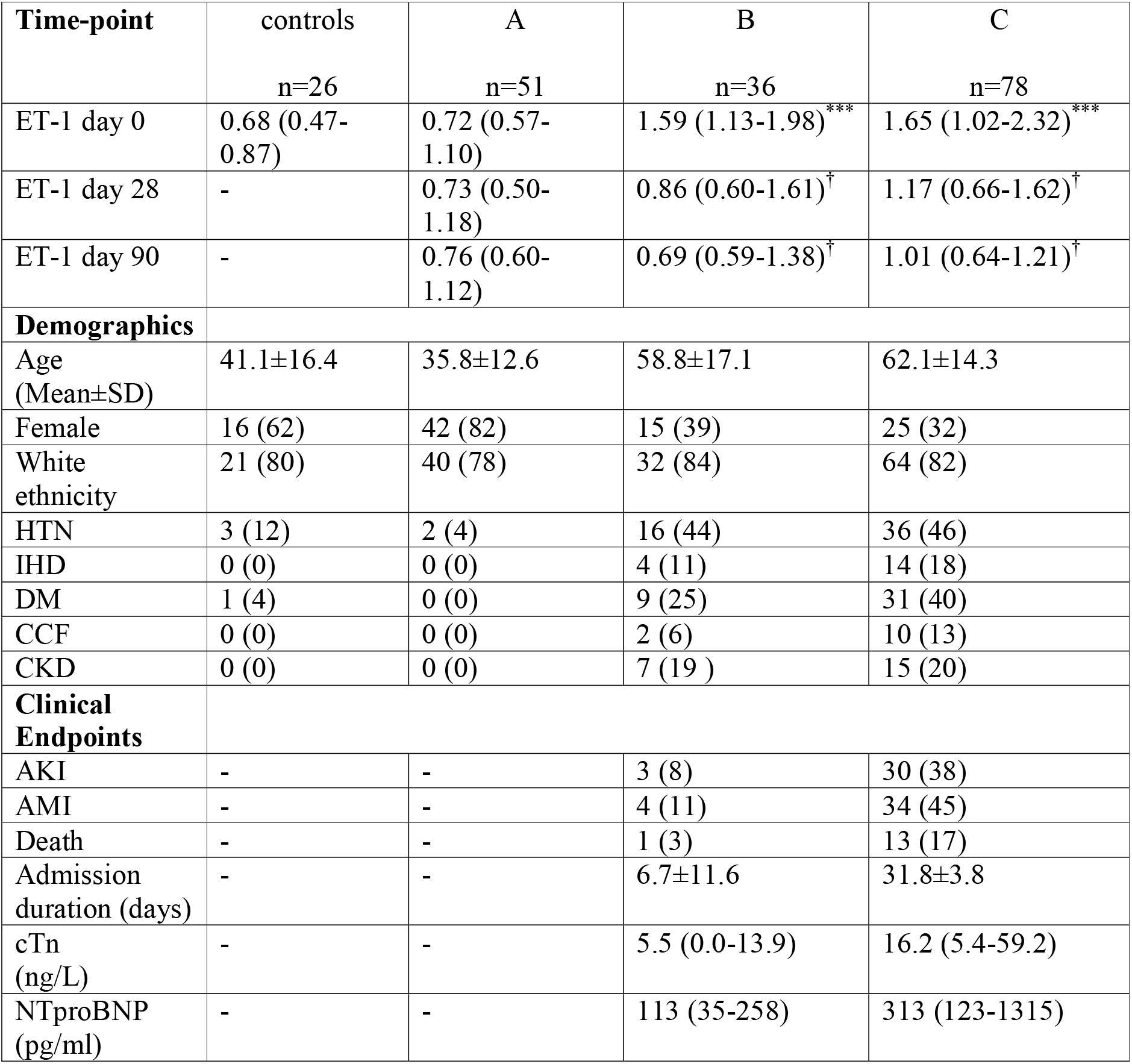
ET-1, patient demographics and clinical endpoints. Table shows: *Top*: ET-1 (pg/ml) at all time-points (median [IQR]). ^***^ indicates significant difference (p=<0.001) when comparing indicated patient group and control; ^†^ indicates significant difference (p=<0.05) when comparing ET-1 at the indicated time-point with the corresponding baseline ET-1 level. *Middle*: Demographics and underlying comorbidities within patient categories, figures are n (% of total). HTN indicates hypertension, IHD: ischemic heart disease, DM: diabetes mellitus, CCF: congestive cardiac failure, CKD: chronic kidney disease. *Bottom*: Clinical endpoints recorded for hospitalised patients, figures are n (% of total). AKI indicates acute kidney injury, AMI: acute myocardial injury. cTn refers to peak cardiac specific Troponin levels (median [IQR]); NTproBNP: peak N-terminal pro-B-type natriuretic peptide (median [IQR]); Admission duration: duration of index Covid-19 related admission measured in total days (mean±SD).

**Figure 1.**
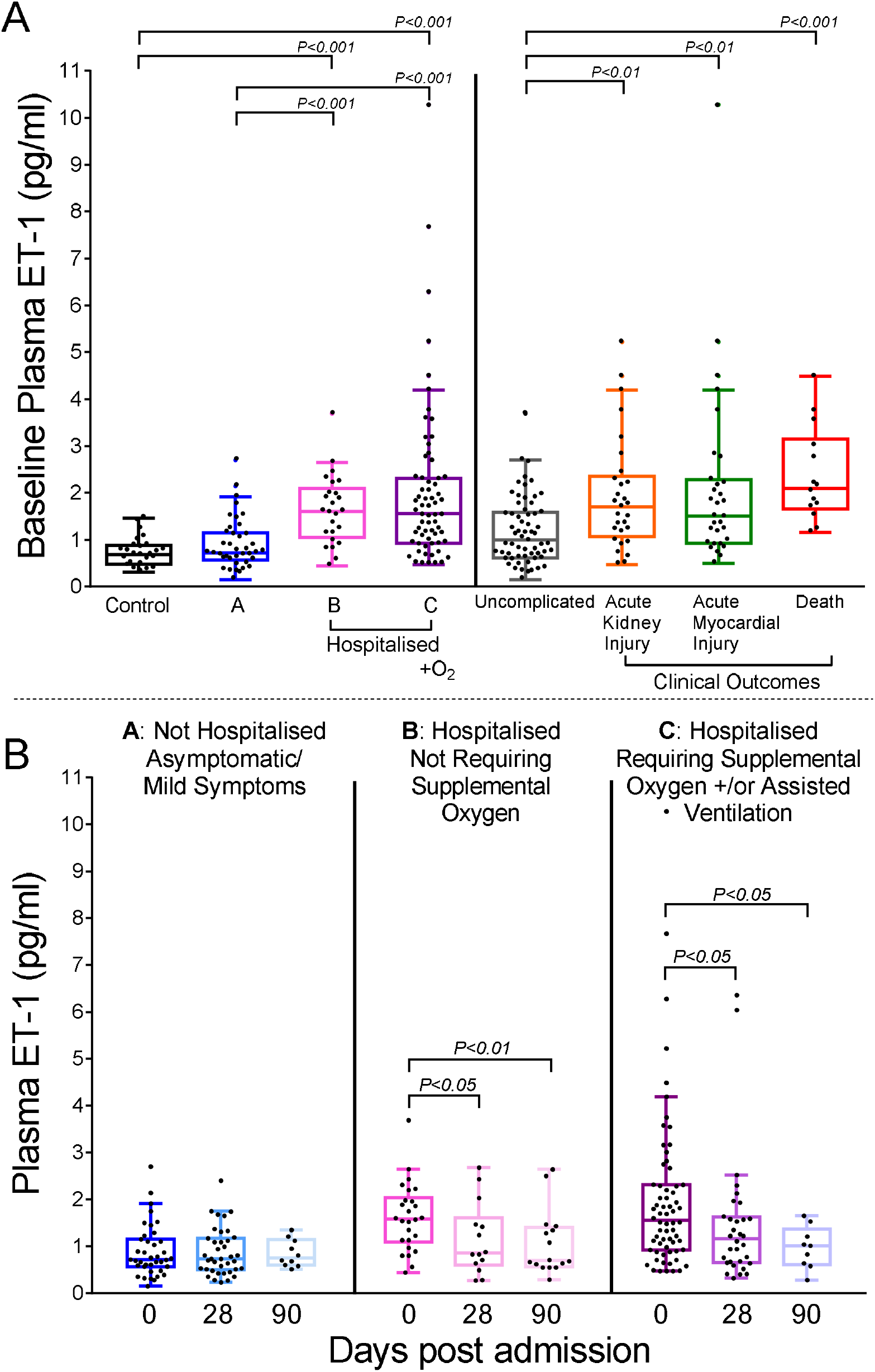
A, Comparison of ET-1 at baseline across categories: Controls refer to non-infected volunteers; group A: non-hospitalised Covid-19 patients; groups B and C: hospitalised Covid-19 patients; uncomplicated: surviving Covid-19 infected patients not requiring supplemental oxygen or assisted ventilation and without acute kidney injury or acute myocardial injury. B, Comparison of ET-1 levels in categories A-C at day 28 and 90 after admission compared to day 0.

Baseline ET-1 levels (pg/ml) were also significantly elevated in subgroups of patients that died (2.09 [1.66-3.15]), developed acute kidney (1.70 [1.07-2.36]) or myocardial injury (1.50 [0.92-2.28]) compared with patients with an uncomplicated infection (1.00 [0.61-1.57], p=<0.01) for whom these endpoints were not recorded (*Figure 1A*).

Amongst surviving hospitalised patients, ET-1 levels (pg/ml) decreased monotonically when measured at 28 days (Group B: 0.86 [0.60-1.61] and Group C: 1.17 [0.66-1.62] versus baseline, p=<0.05) and 90 days (Group B: 0.69 [0.59-1.38] and Group C: 1.01 [0.64-1.21] versus baseline, p=<0.05): *Figure 1B*.

## Discussion

Our results demonstrate increased ET-1 levels in the acute phase of Covid-19 infection in patients requiring hospitalisation. We hypothesise that elevation of ET-1 plasma levels could result from inflammatory cytokine mediated up-regulation of the stimulated Weibel-Palade body ET-1 secretory pathway. Von-Willebrand Factor is the principal stored component of Weibel-Palade bodies^12^ and has been shown to be similarly up-regulated in the acute phase of Covid-19 infection^8^. Alternatively or additionally, viral induced cell death may result in the release of stored ET-1 into the circulation (figure 2).

**Figure 2.**
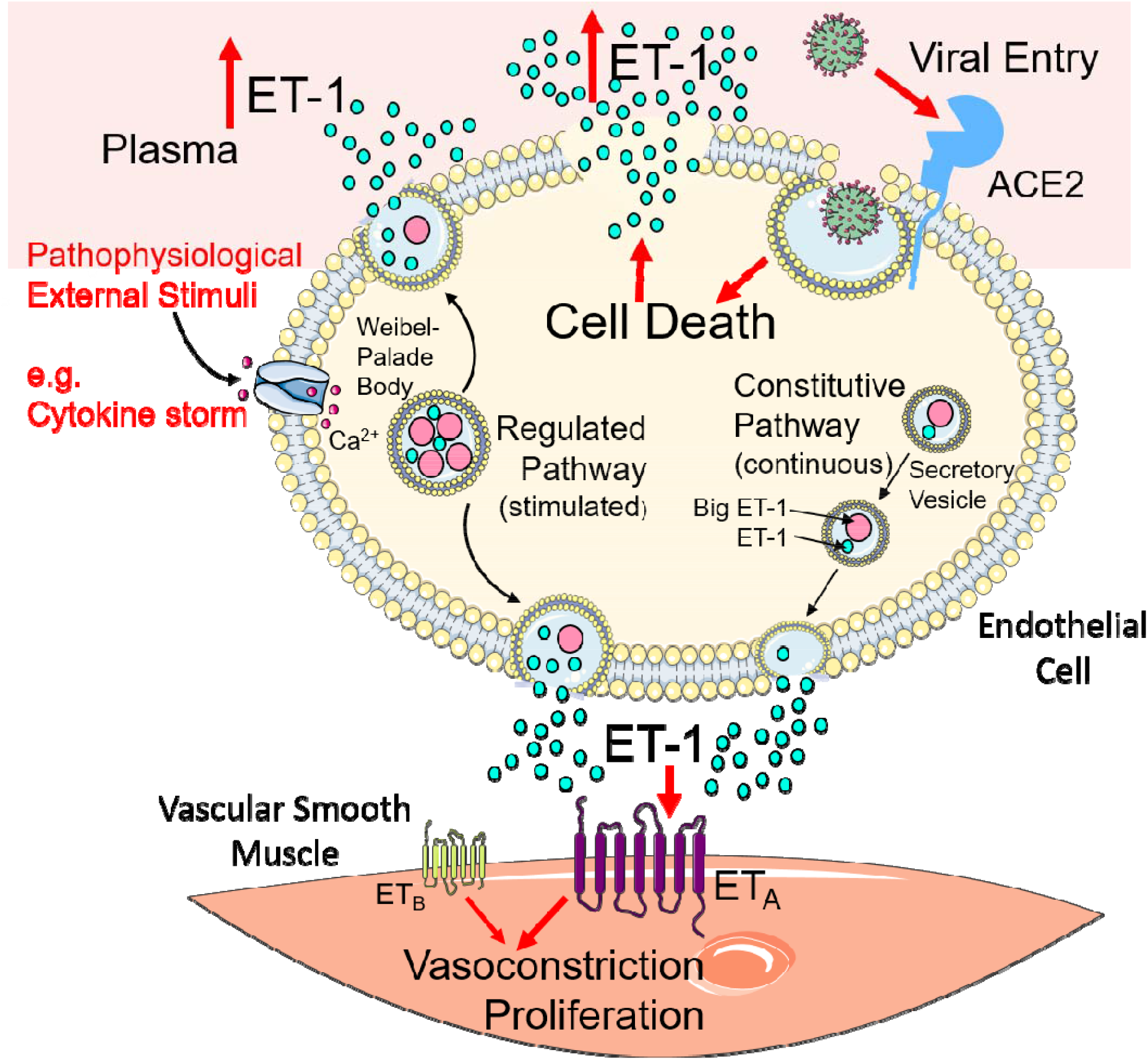
Hypothesis for mechanism of increased plasma ET-1 levels in severe Covid-19 infection. Covid-19 infection may up-regulate circulating cytokines to stimulate the regulated ET-1 secretory pathway. Additionally, viral entry into endothelial cells induces cell damage and release of stored ET-1 into the circulation

Accumulating evidence suggests endothelial dysregulation is a key mechanism for disease progression following Covid-19 infection. Post-mortem histology from Covid-19 patients with multi-organ failure has identified pathological endotheliitis in multiple vascular beds^4,5,6^ while elevation of various biomarkers of endothelial damage in patients with severe Covid-19 infection has also been demonstrated^7,8^.

Previous studies over the last 25 years have shown that increased circulating levels of the endothelial peptide, ET-1 are associated with increased coronary^13^ and systemic vasoconstriction^14^ in coronary microvascular dysfunction, and endothelial dysfunction in vasospastic angina^15^. In our Covid-19 cohort, the highest ET-1 levels were seen in dying patients and those whose infection was complicated by respiratory failure (indicated by a requirement for supplemental oxygen or assisted ventilation), acute kidney or myocardial injury suggesting that increased circulating ET-1 may also be an important contributor to the pathogenesis of these complications.

### Limitations

Our study is limited by its retrospective observational design with all patients enrolled during the first wave of the pandemic, when no specific therapies for Covid-19 were known and prior to the availability of vaccines. Further exploration of the pathophysiological changes resulting from elevated ET-1 levels in critically unwell patients was not feasible in this period.

## Conclusion

Our results demonstrate that elevated ET-1 in the acute phase of Covid-19 infection is a clinical feature unique to severe disease, supporting the hypothesis^8^ that endothelin receptor antagonists, used to treat pulmonary arterial hypertension, may be beneficial in a subset of Covid-19 patients.

## Data Availability

All data that support the findings of this study are available from the corresponding author upon reasonable request

## Acknowledgements

We thank all participating patients and volunteers for their contribution to this research and the staff of NIHR BioResource for Translational Research for collection and processing of plasma samples.

## Note to Referees/supplemental information

All data that support the findings of this study are available from the corresponding author upon request. To assist the review process, we have provided below further details of the statistical analyses performed as well as the following details of the possible confounding variables considered in this study and Univariate Analyses of Co-variance.

All statistical analysis was performed using SPSS version 27 (IBM Corp., USA) for Windows. Normality of continuous variable distributions was tested by one-sample Kolmogorov-Smirnov test. Non-normally distributed continuous variables are presented as Median (inter-quartile range [Q1 - Q3]). Due to the skewed distribution of ET-1, no potential or extreme outliers have been excluded from analysis. Outliers were noted to mostly occur in patient category C (hospitalised and requiring supplemental oxygen/assisted ventilation) or amongst subgroups with clinical complications of Covid-19 infection therefore we have assumed these values reflect true biological variability in the study population. Comparison of ET-1 between patient categories was performed using the Independent Samples Kruskal-Wallis Test and also between time-points for each patient category. In the latter case, an independent samples non-parametric test was chosen to increase power given the low number of paired corresponding samples available due to patient drop-out at 28 and 90 days. Reasons for loss of patients to follow up were in many cases unavoidable including patient death, persisting disability and patients being repatriated outside our local area having been referred to our centre for specialist tertiary care during their initial illness.

### Demographic details of patient categories and univariate analyses of co-variance

Covariate variables are presented as n (%) unless otherwise stated. No demographic or clinical endpoint data was available for 3/39 patients in category B otherwise clinical variables were available for all other enrolled patients. Test statistic: difference in mean square ET-1 at time 0 between patient categories (controls, A, B and C) after adjustment for covariate variables; p value after univariate adjustment refers to probability that differences in baseline ET-1 are not significantly different between patient categories (controls, A, B and C) after adjustment for differences in covariate variables.

| Covariate Variables | controls<br>n=26 | A<br>n=51 | B<br>n=36 | C<br>n=78 | Test<br>statistic | p value after univariate<br>adjustment |
| --- | --- | --- | --- | --- | --- | --- |
| Age<br>(Mean±SD) | 41.1±16.4 | 35.8±12.6 | 58.8±17.1 | 62.1±14.3 | 2.91 | 0.04 |
| Female | 16 (62) | 42 (82) | 15 (39) | 25 (32) | 13.4 | <0.001 |
| HTN | 3 (12) | 2 (4) | 16 (44) | 36 (46) | 7.3 | <0.001 |
| IHD | 0 (0) | 0 (0) | 4 (11) | 14 (18) | 8.9 | <0.001 |
| DM | 1 (4) | 0 (0) | 9 (25) | 31 (40) | 7.5 | <0.001 |
| CCF | 0 (0) | 0 (0) | 2 (6) | 10 (13) | 7.8 | <0.001 |
| CKD | 0 (0) | 0 (0) | 7 (19) | 15 (20) | 9.0 | <0.001 |

## Notes

Funding: Supported in full or part by The Jon Moulton Charity Trust (GRA, APD, SPH); National Institute of Health Research, Cambridge Biomedical Research Centre, BRC-1215-20014 (JC); Biomedical Resources Grant, University of Cambridge, Cardiovascular Theme RG64226 (GRA, REK, APD), British Heart Foundation, TG/18/4/33770, (APD, JJM). The views expressed are those of the author(s) and not necessarily those of the NIHR or the Department of Health and Social Care.

### Competing Interest Statement

MA, PJG and PA are employees of AstraZeneca (UK) Plc. All other authors have no disclosures.

### Funding Statement

The study was funded in full or part by The Jon Moulton Charity Trust (GRA, APD, SPH); National Institute of Health Research, Cambridge Biomedical Research Centre, BRC-1215-20014 (JC); Biomedical Resources Grant, University of Cambridge, Cardiovascular Theme RG64226 (GRA, REK, APD), British Heart Foundation, TG/18/4/33770, (APD, JJM). The views expressed are those of the author(s) and not necessarily those of the NIHR or the Department of Health and Social Care.

### Author Declarations

Ethical approval was obtained from the East of England: Cambridge Central Research Ethics Committee ('NIHR Bioresource': REC:17/EE/0025)

## REFERENCES

1. Alexander SPH, Armstrong JF, Davenport AP, et al. A rational roadmap for SARS-CoV-2/COVID-19 pharmacotherapeutic research and development: IUPHAR Review 29. Br J Pharmacol. 2020;177(21):4942–4966.

2. Calabretta E, Moraleda JM, Iacobelli M, et al. COVID-19-induced endotheliitis: emerging evidence and possible therapeutic strategies. Br J Haematol. 2021;193(1):43–51.

3. Fisk M, Althage M, Moosmang S, et al. Endothelin antagonism and sodium glucose Co-transporter 2 inhibition. A potential combination therapeutic strategy for COVID-19. Pulm Pharmacol Ther. 2021;69:102035.

4. Ackermann M, Verleden SE, Kuehnel M, et al. Pulmonary Vascular Endothelialitis, Thrombosis, and Angiogenesis in Covid-19. N Engl J Med. 2020;383(2):120–128.

5. Varga Z, Flammer AJ, Steiger P, et al. Endothelial cell infection and endotheliitis in COVID-19. Lancet. 2020;395(10234):1417–1418.

6. Fox SE, Falgout L, Vander Heide RS. COVID-19 myocarditis: quantitative analysis of the inflammatory infiltrate and a proposed mechanism. Cardiovasc Pathol. 2021;54:107361.

7. Thwaites RS, Sanchez Sevilla Uruchurtu A, Siggins MK, et al. Inflammatory profiles across the spectrum of disease reveal a distinct role for GM-CSF in severe COVID-19. Sci Immunol. 2021;6(57):eabg9873.

8. Goshua G, Pine AB, Meizlish ML, et al. Endotheliopathy in COVID-19-associated coagulopathy: evidence from a single-centre, cross-sectional study. Lancet Haematol. 2020;7(8):e575–e582.

9. Davenport AP, Hyndman KA, Dhaun N, et al. Endothelin. Pharmacol Rev. 2016;68(2):357–418.

10. Russell FD, Davenport AP. Secretory pathways in endothelin synthesis. Br J Pharmacol. 1999;126(2):391–398.

11. Thenappan T, Ormiston ML, Ryan JJ, Archer SL. Pulmonary arterial hypertension: pathogenesis and clinical management. BMJ. 2018;360:j5492. Published 2018 Mar 14.

12. Rondaij MG, Bierings R, Kragt A, van Mourik JA, Voorberg J. Dynamics and plasticity of Weibel-Palade bodies in endothelial cells. Arterioscler Thromb Vasc Biol. 2006;26(5):1002–1007.

13. Cox ID, Bøtker HE, Bagger JP, Sonne HS, Kristensen BO, Kaski JC. Elevated endothelin concentrations are associated with reduced coronary vasomotor responses in patients with chest pain and normal coronary arteriograms. J Am Coll Cardiol. 1999;34(2):455–460.

14. Ford TJ, Corcoran D, Padmanabhan S, et al. Genetic dysregulation of endothelin-1 is implicated in coronary microvascular dysfunction. Eur Heart J. 2020;41(34):3239–3252. doi:10.1093/eurheartj/ehz915

15. Lerman A, Holmes DR Jr, Bell MR, Garratt KN, Nishimura RA, Burnett JC Jr. Endothelin in coronary endothelial dysfunction and early atherosclerosis in humans. Circulation. 1995;92(9):2426–2431.

